# Optimizing Ocular Pathology Classification with CNNs and OCT Imaging: A Systematic and Performance Review

**DOI:** 10.1101/2024.06.18.24309070

**Authors:** Walter Hauri-Rosales, Oswaldo Pérez, Marlon Garcia-Roa, Ellery López-Star, Ulises Olivares-Pinto

**Author notes:** Corresponding author ✉ (U. Olivares-Pinto).

## Abstract

Vision loss due to chronic-degenerative diseases is a primary cause of blindness worldwide. Deep learning architectures utilizing optical coherence tomography images have proven effective for the early diagnosis of ocular pathologies. Nevertheless, most studies have emphasized the best outcomes using optimal hyperparameter combinations and extensive data availability. This focus has eclipsed the exploration of how model learning capacity varies with different data volumes. The current study evaluates the learning capabilities of efficient deep-learning classification models across various data amounts, aiming to determine the necessary data portion for effective clinical trial classifications of ocular pathologies. A comprehensive review was conducted, which included 295 papers that employed OCT images to classify one or more of the following retinal pathologies: Drusen, Diabetic Macular Edema, and Choroidal Neovascularization. Performance metrics and dataset details were extracted from these studies. Four Convolutional Neural Networks were selected and trained using three strategies: initializing with random weights, fine-tuning, and retraining only the classification layers. The resultant performance was compared based on training size and strategy to identify the optimal combination of model size, dataset size, and training approach. The findings revealed that, among the models trained with various strategies and data volumes, three achieved 99.9% accuracy, precision, recall, and F1 score. Two of these models were fine-tuned, and one used random weight initialization. Remarkably, two models reached 99% accuracy using only 10% of the original training dataset. Additionally, a model that was less than 10% the size of the others achieved 98.7% accuracy and an F1 score on the test set while requiring 100 times less computing time. This study is the first to assess the impact of training data size and model complexity on performance metrics across three scenarios: random weights initialization, fine-tuning, and retraining classification layers only, specifically utilizing optical coherence tomography images.

## 1. **Introduction**

Chronic-degenerative diseases are a leading cause of death globally, accounting for 70% of all deaths. According to the World Health Organization (WHO), around 422 million individuals are affected by Type 1 and 2 Diabetes Mellitus (DM) [1]. The International Diabetes Federation (IDF) projected that this number could rise to 700 million by 2045 [2]. In 2020, DM ranked as the ninth leading cause of death worldwide [3]. A long-term consequence of inadequate glycemic control in DM patients is visual impairment, which may lead to various ocular pathologies [4], [5], [6].

Among these, Age-related Macular Degeneration (AMD), a major cause of blindness globally, affects millions and is particularly severe in individuals over 60 in developed countries. It is categorized into two types: neovascular (wet) and non-neovascular (dry). Dry AMD, which accounts for 85% of cases, generally has a more favorable prognosis compared to wet AMD, responsible for about 80% of severe vision loss from the condition [7], [8]. Consequences of poor vision include increased risk of falls, depression, and the need for long-term care if unable to perform activities of daily living, such as dressing, eating, and working.

Diabetic Retinopathy (DR), prevalent in approximately 33% of DM patients, leads the causes of preventable blindness worldwide [2], [9]. Other related conditions include Diabetic Macular Edema (DME), Drusen formation, and Choroidal neovascularization (CNV). This study focuses on analyzing classification methods to detect these three pathologies using medical imaging [10] [11]. Recent advances have shown that deep learning algorithms, particularly using optical coherence tomography (OCT) [12], [13], [14] and fundus images [15], [16], can automatically extract pathological features. Furthermore, features identified in one pathology may be applicable to others, allowing for efficient classification across multiple conditions [17].

Convolutional Neural Networks (CNNs), first proposed by Krizhevsky, are effective for extracting features from medical images [18]. A typical CNN model includes several layers: convolutional layers that detect complex patterns, a nonlinear activation function, and a pooling layer that reduces dimensionality to enhance model robustness against small changes in input. The process concludes with fully connected layers that lead to a classification output. The training of CNNs involves adjusting internal weights to minimize a cost function, followed by a transfer learning process to apply learned features to new datasets.

This paper assesses various deep learning architectures for detecting retinal pathologies, comparing performance metrics against the current state of the art. It also evaluates the impact of training data volume, model complexity, and their influence on performance in scenarios including random weight initialization, fine-tuning through transfer learning, and retraining only the classification layers. Despite most studies achieving over 99% accuracy in classifying retinal pathologies with OCT images [11], [14], [19], [20], [21], [22], [23],[24] the relationship between model learning capacity and data volume has been underexplored. This study aims to address this gap, potentially guiding the data requirements for clinical trials to effectively classify ocular pathologies.

## 2. **Materials and Methods**

This study evaluated the capability of recent CNNs architectures to detect retinal pathologies using Optical Coherence Tomography images. The assessment was divided into two phases: first, a systematic review of current deep learning classification methods was conducted; second, the performance of the most effective CNNs, trained to differentiate between normal and three specific retinal pathologies, was compared.

### 2.1. **Systematic Review of Classification Algorithms**

A comprehensive literature survey was performed using three journal databases: Web of Science, PubMed, and IEEE Xplore. The objective was to collect prior studies involving OCT images and deep learning. The search was structured around the following query: (OCT **OR** “Optical Coherence Tomography”) **AND** (Retina) **AND** (Classification) **AND** (“Deep Learning” **OR** “Convolutional Neural Network” **OR** “CNN” **OR** “Machine Learning”).

The query illustrated in Figure 1 outlines the systematic process of identifying, screening, and including studies for a review on the application of deep learning models for classifying retinal pathologies using Optical Coherence Tomography (OCT) images, based on the PRISMA methodology {Citation}. Initially, records were identified from three databases: IEEE (n = 204), Web of Science (n = 242), and PubMed (n = 224), resulting in a total of 670 records. Prior to screening, 118 duplicate records and 443 records marked as ineligible by automation tools were removed, leaving 109 records for screening. During the screening phase, all 109 records were reviewed for relevance, and no records were excluded or not retrieved. In the eligibility assessment stage, the 109 reports were evaluated based on the inclusion criteria, leading to the exclusion of 40 reports that did not include the pathology of interest and 14 reports that did not utilize deep learning models. Ultimately, 55 new studies were included in the final review, with no additional reports of new included studies. This thorough and unbiased selection process ensured that only studies relevant to the research question were included, focusing on the application of deep learning techniques in OCT image analysis for retinal pathologies. The comparative outcomes of these studies are detailed in Table 1.

**Figure 1:**
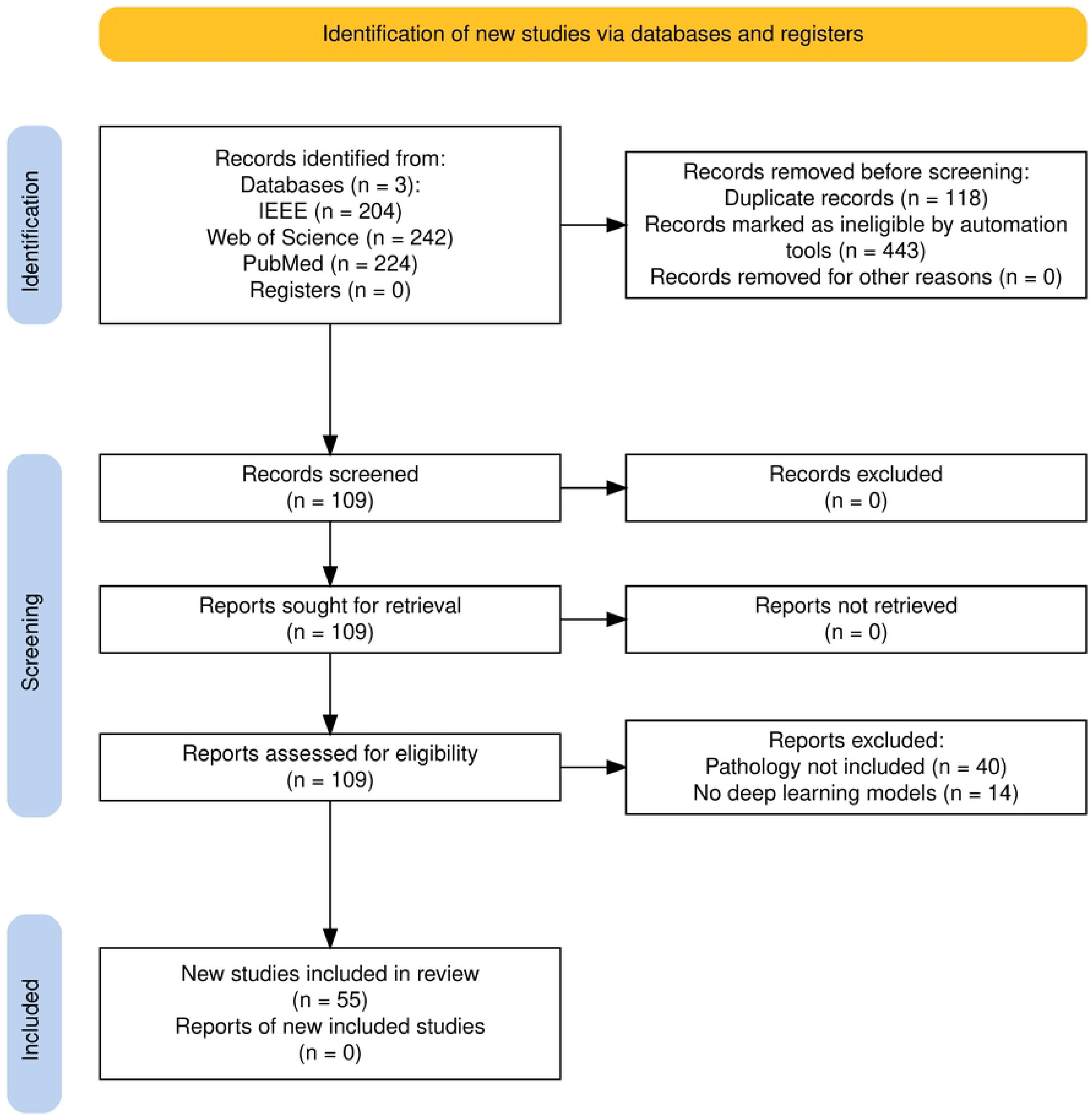
Paper Query Results and Selection Process. From the three journal databases, we initially retrieved 358 papers. After removing duplicates, 295 papers remained. We then conducted a refined query to identify papers that discussed at least one of the specific pathologies, narrowing it down to 64 papers. Ultimately, 43 of these papers, which utilized machine learning-based classification techniques, were selected for further analysis.

**Table 1:**
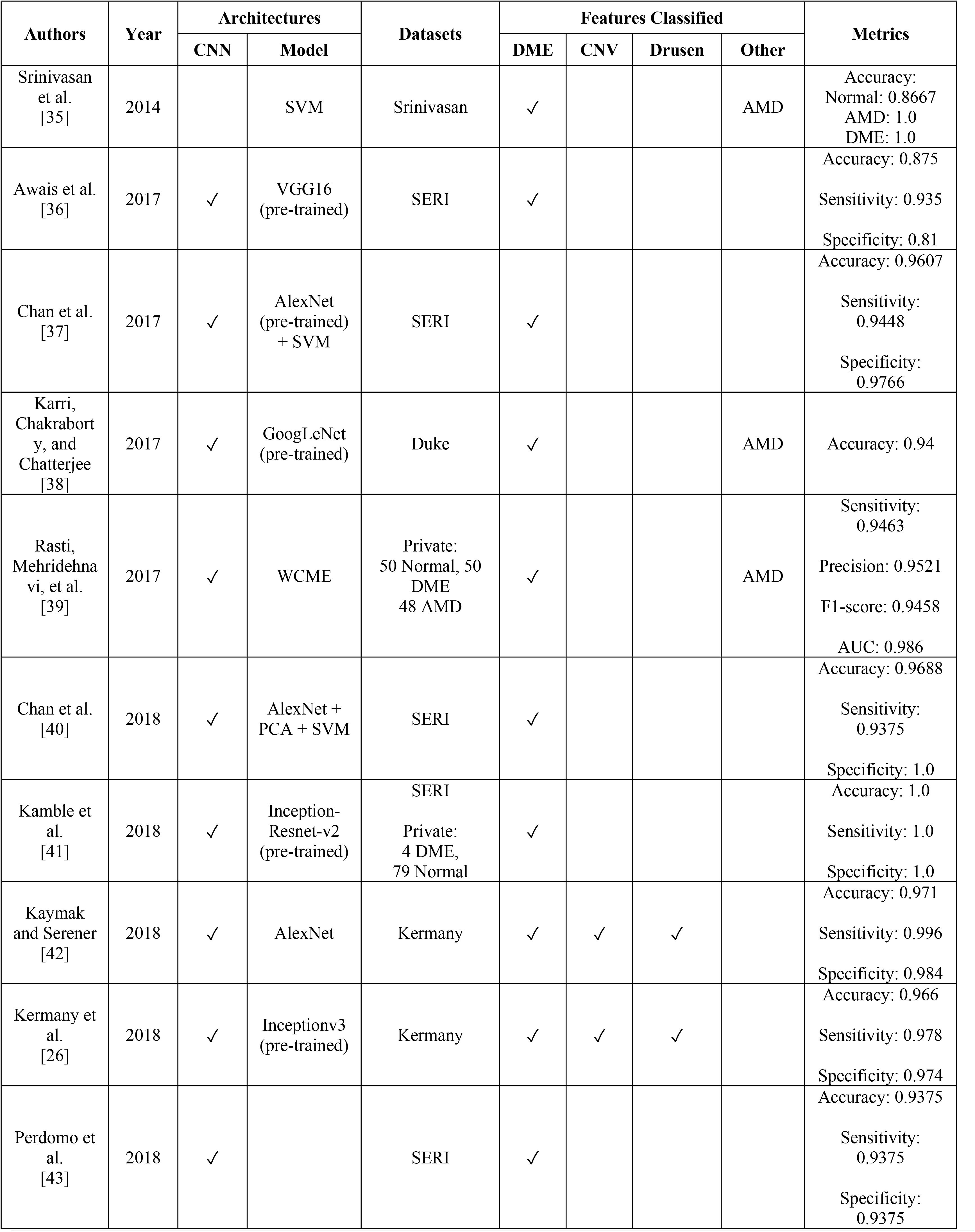
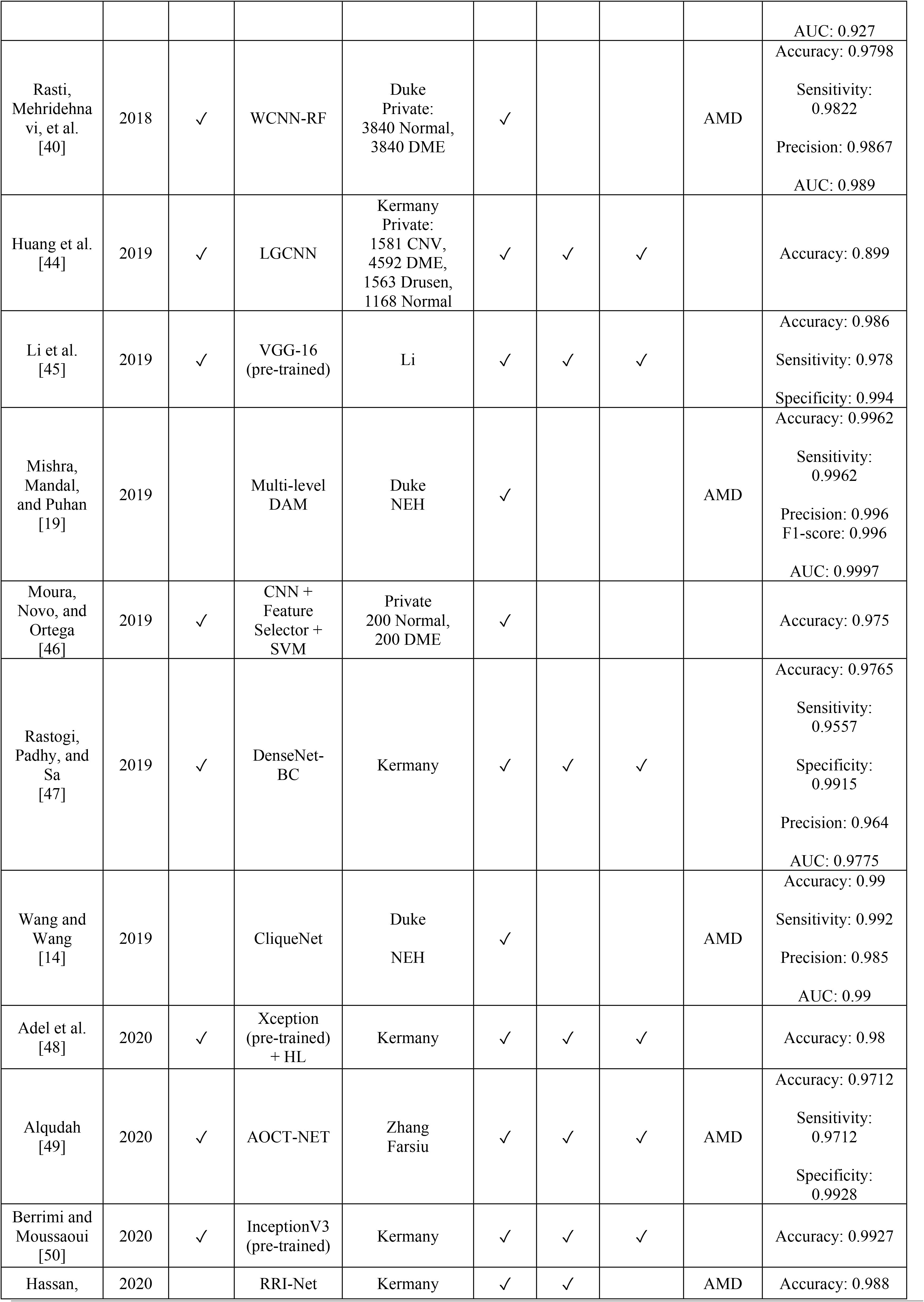
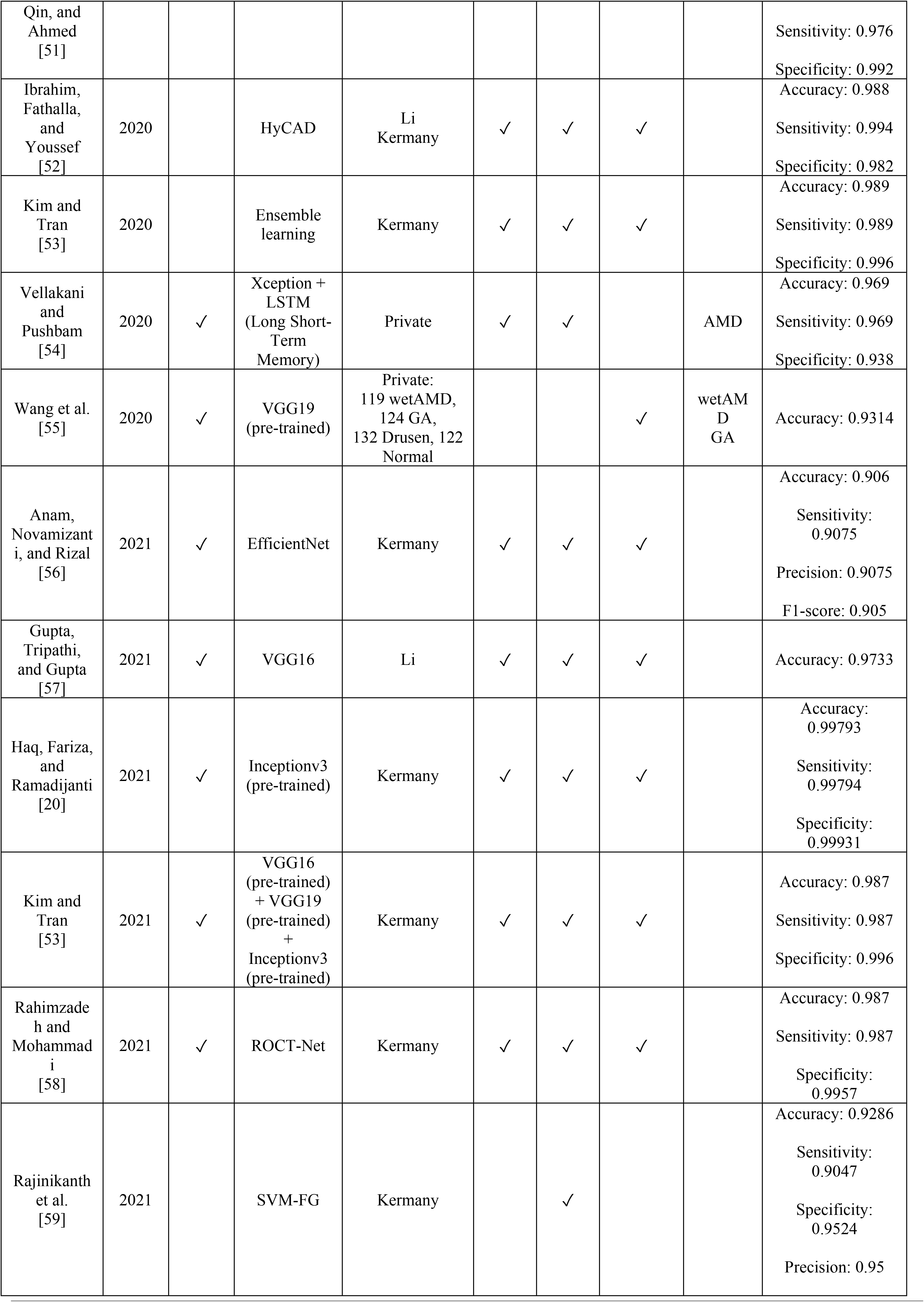
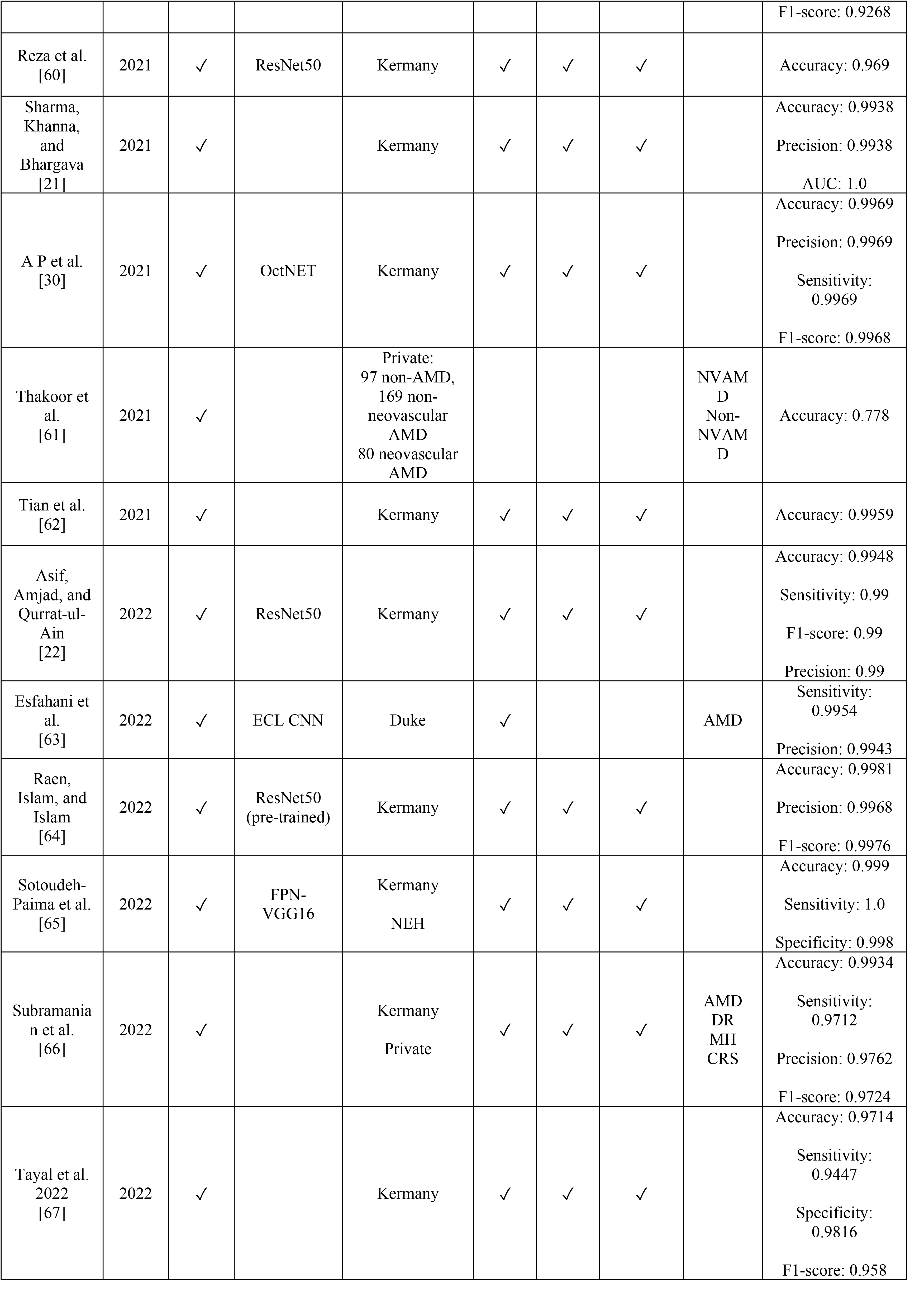
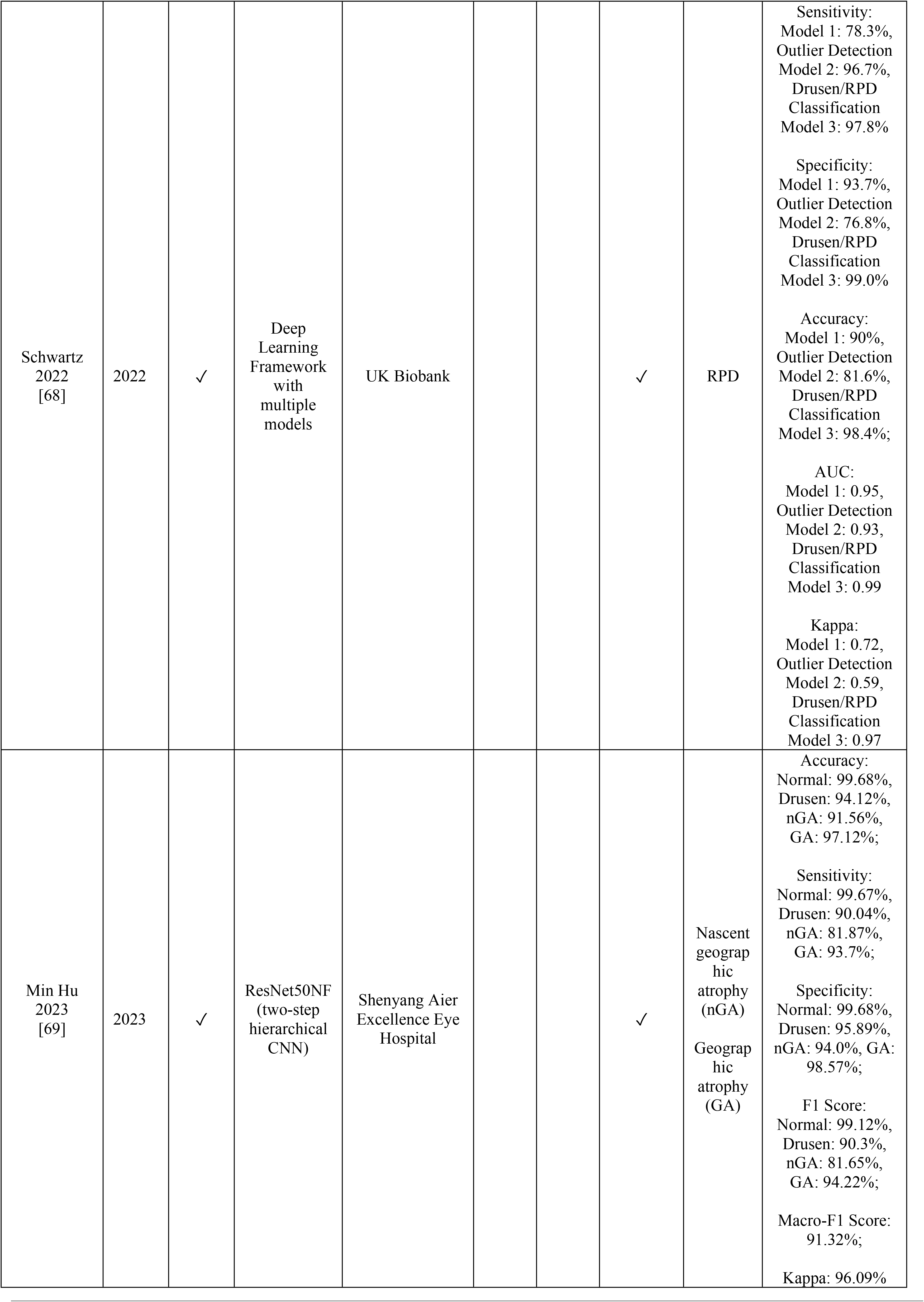
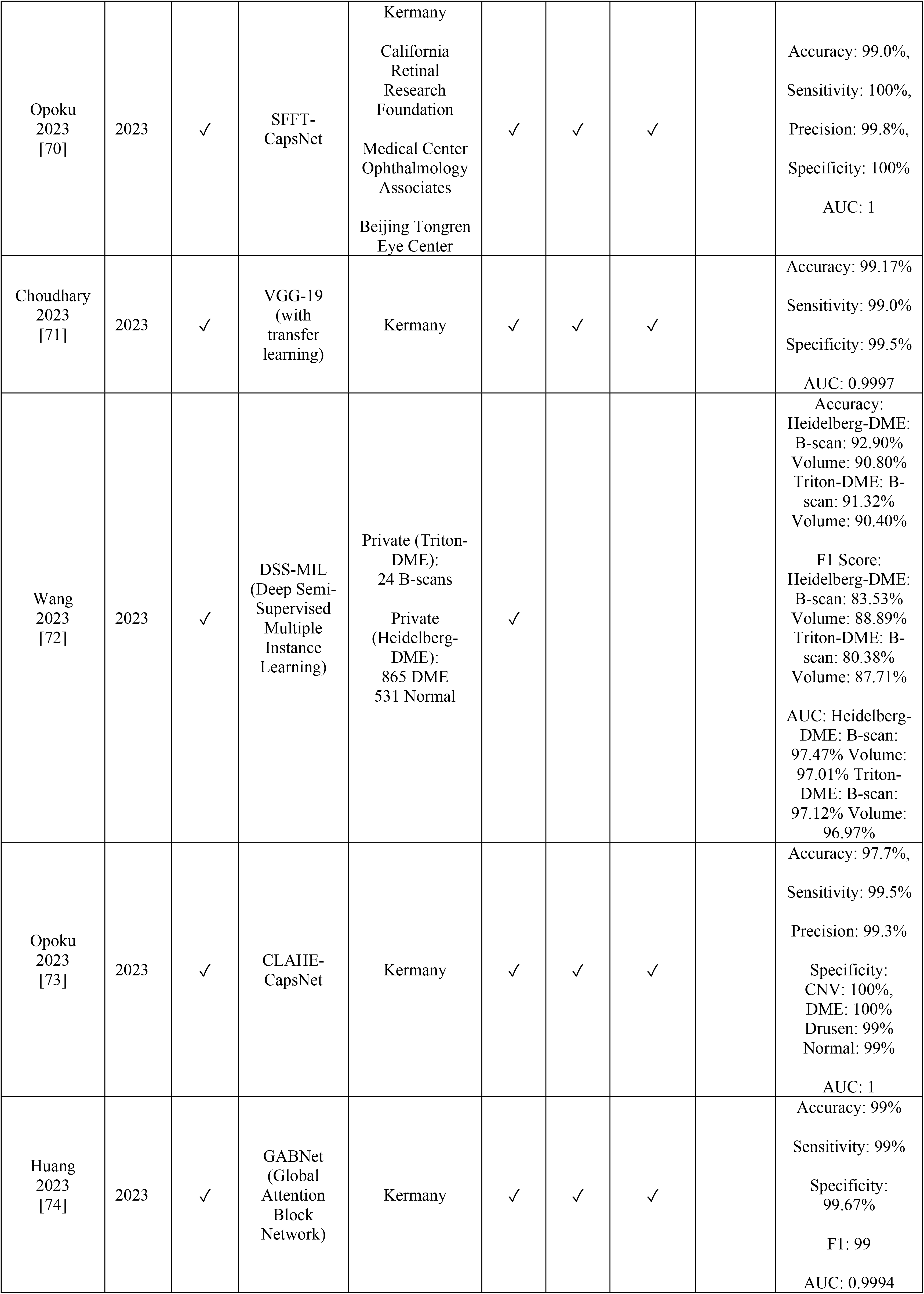
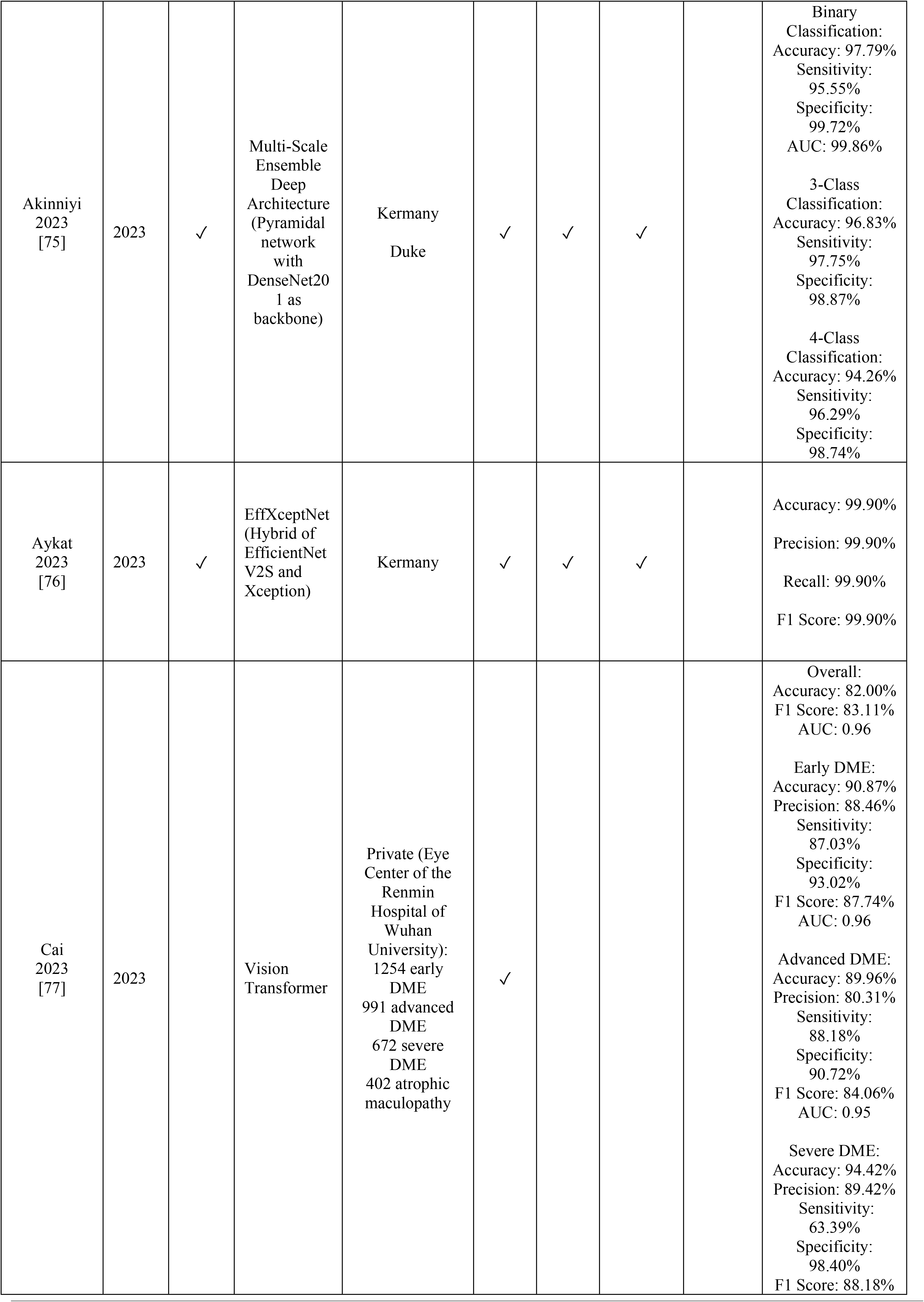
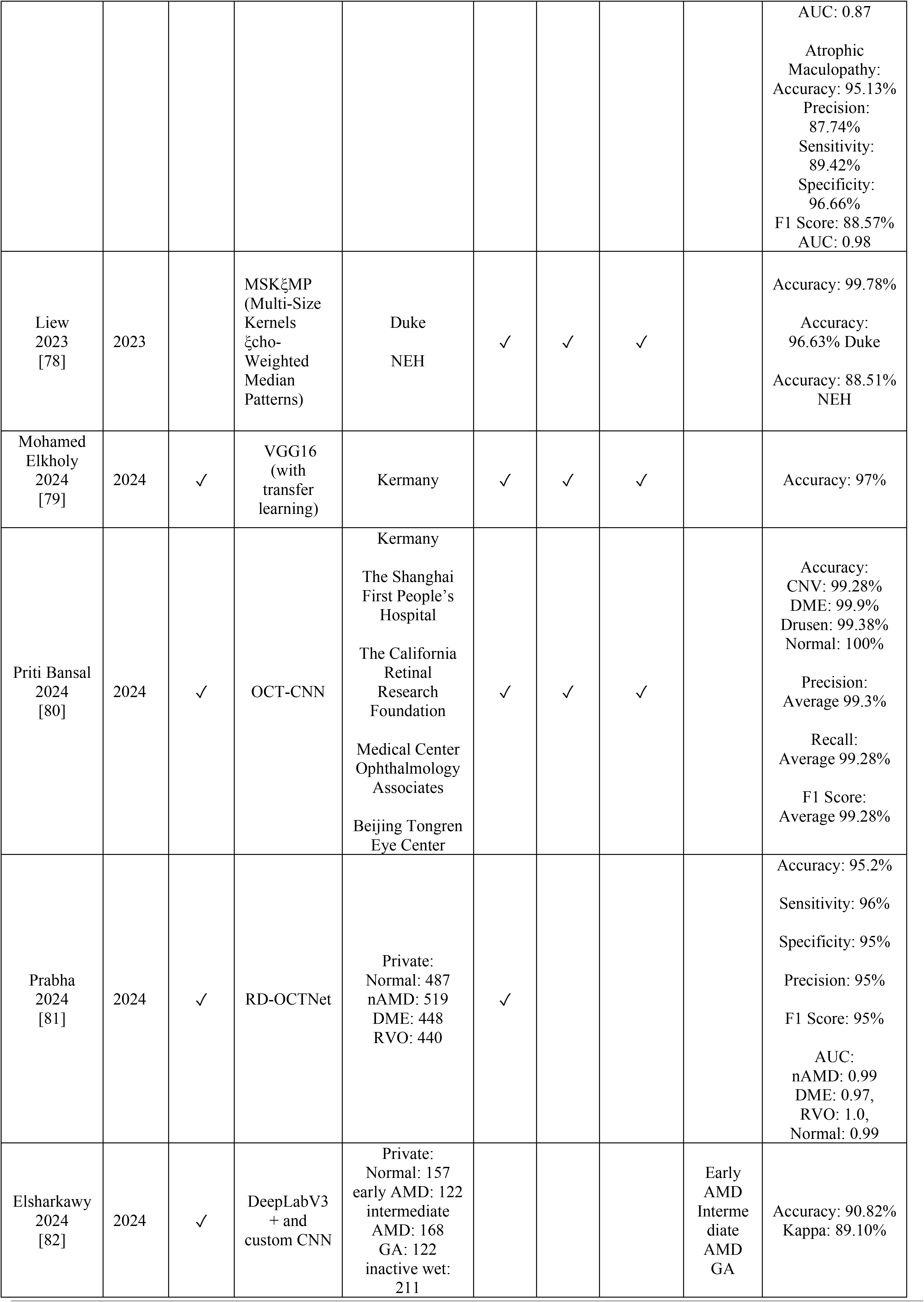
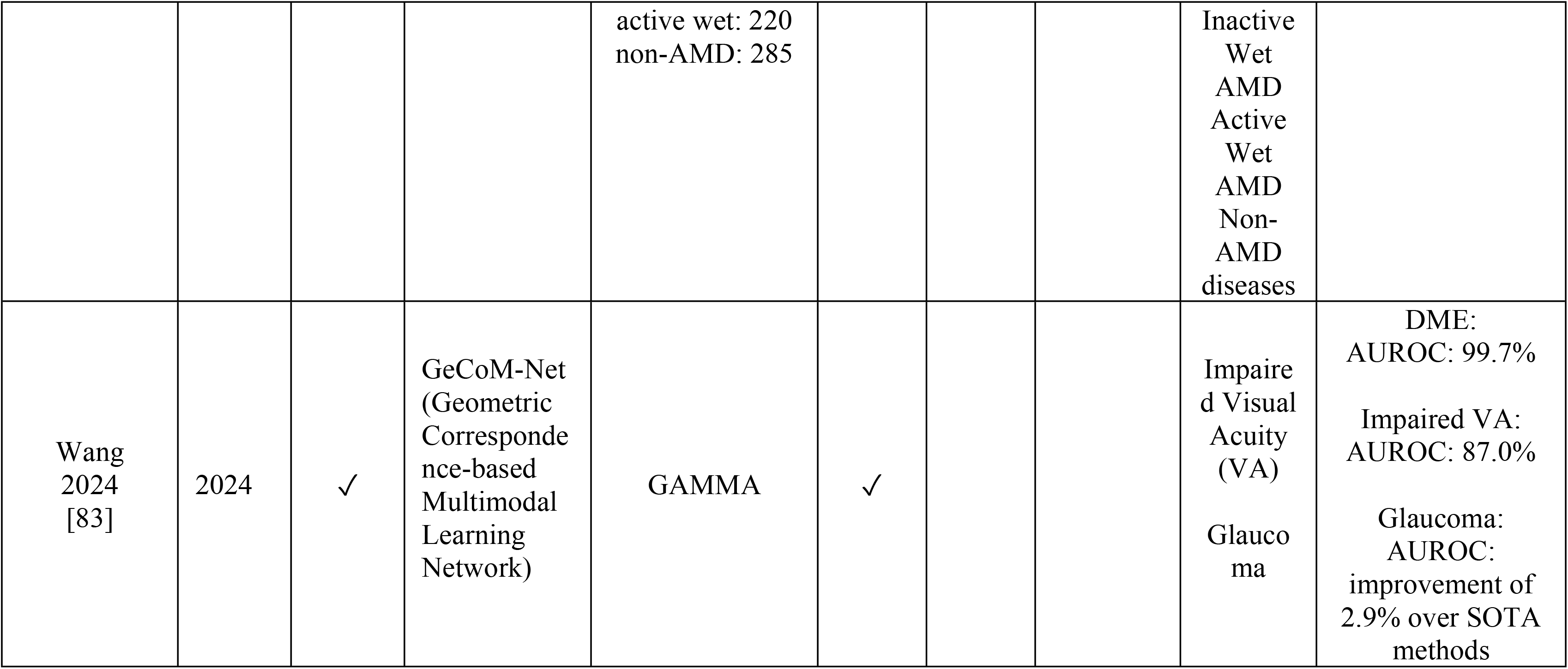
Summary of Studies on Deep Learning Architectures for OCT Image-Based Retinal Pathology Classification, Detailing Authors, Publication Years, CNN Architectures, Datasets, Classified Pathologies, and Performance Metrics.

The trend shows an increasing annual publication rate in this field. The utilization of pre-trained CNNs has emerged as a predominant method for classifying retinal pathologies from OCT images, with some studies introducing minor modifications to these architectures to enhance performance. The employment of such pre-trained models stands out as one of the most common and practical strategies in the literature. Regarding datasets, most of the papers selected via the methodology outlined above relied on publicly available datasets, though several studies also incorporated privately sourced images. Table 2 presents a compilation of these public datasets, including citations and the distribution of data used in each study. Notably, the dataset provided by Kermany et al. [26] is frequently cited and utilized for training and testing deep learning classifiers. This dataset encompasses images associated with three retinal pathologies: DME, CNV, and Drusen.

**Table 2:** Overview of public OCT datasets highlighting image counts across different retinal pathologies.

| **Dataset Source** | **Normal** | **Dry AMD** | **DME** | **CNV** | **Drusen** |
| --- | --- | --- | --- | --- | --- |
| Duke/Srinivasan [35] | 15 | 15 | 15 | 0 | 0 |
| SERI [84] | 64 | 0 | 64 | 0 | 0 |
| Kermany [26] | 26,315 | 0 | 11,348 | 37,205 | 8,616 |
| Li et al. [45] | 51,390 | 0 | 11,599 | 37,456 | 8,867 |
| Rasti et al. [40] | 2,375 | 0 | 1,185 | 0 | 1,524 |

### 2.2. **Performance Review of Deep Learning Architectures**

Having identified the most effective classification methods from the literature, as determined by their performance metrics on the Kermany OCT dataset, this study aims to compare the performance of these methods. It will also assess the influence of model learning capacity when exposed to varying amounts of input data. This evaluation will be conducted under three distinct scenarios. The following subsections provide a detailed description of this comparative analysis process.

#### 2.2.1. **Top-Ranking Models**

In this study, we selected four distinct models for an in-depth performance comparison in classifying retinal pathologies using the OCT dataset mentioned earlier. The selected models are:

- Xception: Introduced in 2015 with updates in 2017, known for its depth and complexity [27].
- ResNet-50: A convolutional neural network (CNN) proposed in 2016, recognized for its deep residual learning framework [28].
- OpticNet: The leading CNN for DME classification, first introduced in 2019 [29].
- OctNET: A novel and efficient approach for retinal disease classification, introduced in 2021 [30].

OpticNet and OctNET were specifically pre-trained on the Kermany OCT dataset, while Xception and ResNet-50 were pre-trained on the ImageNet dataset, a large visual database, which contains more than 14 million images hand-annotated [31]. Prior studies have shown that the use of pre-trained models can enhance the learning process by leveraging previously learned features to recognize basic shapes, which can be beneficial for medical imaging tasks [32], [33].

#### 2.2.2. **Dataset for Training Models**

The widely recognized Kermany et al. dataset is extensively used for training and testing models in retinal pathology classification. Hosting 84,484 images, it categorizes them into four classes: Normal, CNV, Drusen, and DME. The dataset is divided into training, testing, and validation subsets. The allocation of images to each subset is detailed in Table 3. The dataset is publicly available and can be accessed online for research purposes [26].

**Table 3:** Distribution of images in the Kermany dataset across training, testing, and validation sets by category.

| **Label** | **Train** | **Test** | **Validation** |
| --- | --- | --- | --- |
| NORMAL | 17002 | 5313 | 4250 |
| CNV | 23971 | 7491 | 5993 |
| DME | 7423 | 2320 | 1855 |
| DRUSEN | 5674 | 1773 | 1419 |

#### 2.2.3. **Training Comparison**

To compare training effectiveness, this study evaluated model performance by partitioning the entire dataset of 84,484 images into smaller, incremental subsets representing specific percentages of the total dataset: 1%, 2.5%, 5%, 7.5%, 9%, 10%, 20%, 40%, 60%, 75%, 90%, and 100%. Each subset was further divided, allocating 64% of the images for training, 20% for testing, and 16% for validation. Each model underwent training with these subsets and was assessed against the consistent validation and test sets established earlier. The selection of training images was randomized for each subset size but fixed across models for comparative consistency.

##### Hyperparameters

During the training phase, all models were trained for 30 epochs with batches of 50 images each. For OpticNet, OctNET, and Xception, the ADAM optimizer was employed with a learning rate set at 0.001. Conversely, ResNet-50 demonstrated improved performance using the Stochastic Gradient Descent with Momentum (SGDM) optimizer, utilizing a learning rate of 0.1 and momentum of 0.6. The specific parameter configurations and outcomes for each CCN model are detailed in Table 4.

**Table 4:**
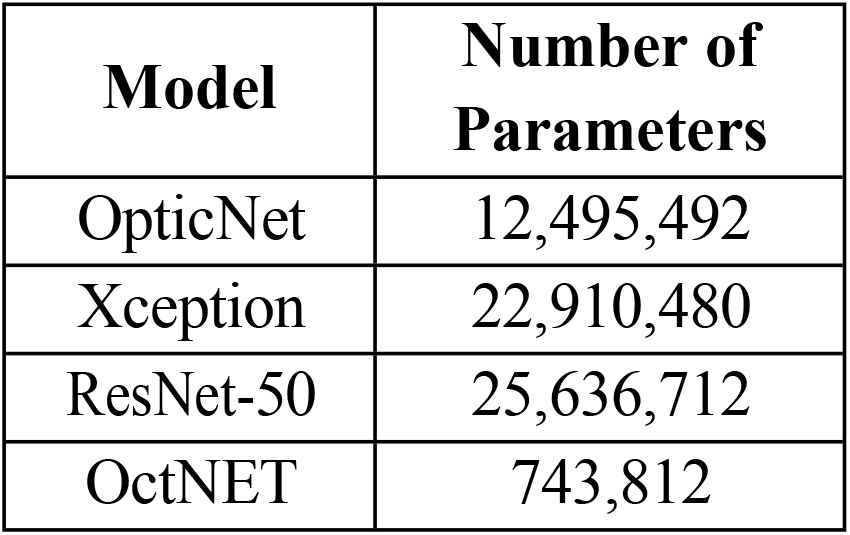
Number of trainable parameters for each evaluated model, reflecting their complexity.

| **Model** | **Number of Parameters** |
| --- | --- |
| OpticNet | 12,495,492 |
| Xception | 22,910,480 |
| ResNet-50 | 25,636,712 |
| OctNET | 743,812 |

##### Training Methodology

The training methodology involved repeating the process across three different model states, as outlined in Table 5, and depicted in Figure 2, to assess the efficacy of model training. The approach is codified in the Algorithm 1, utilizing nested loops; the outermost loop cycles through the model states, the next iterates over varying percentages of the OCT image dataset, and the innermost loop conducts training on each architecture variant with the allocated image subsets. Performance metrics for each model were recorded against the testing set upon completion of training.

**Figure 2:**
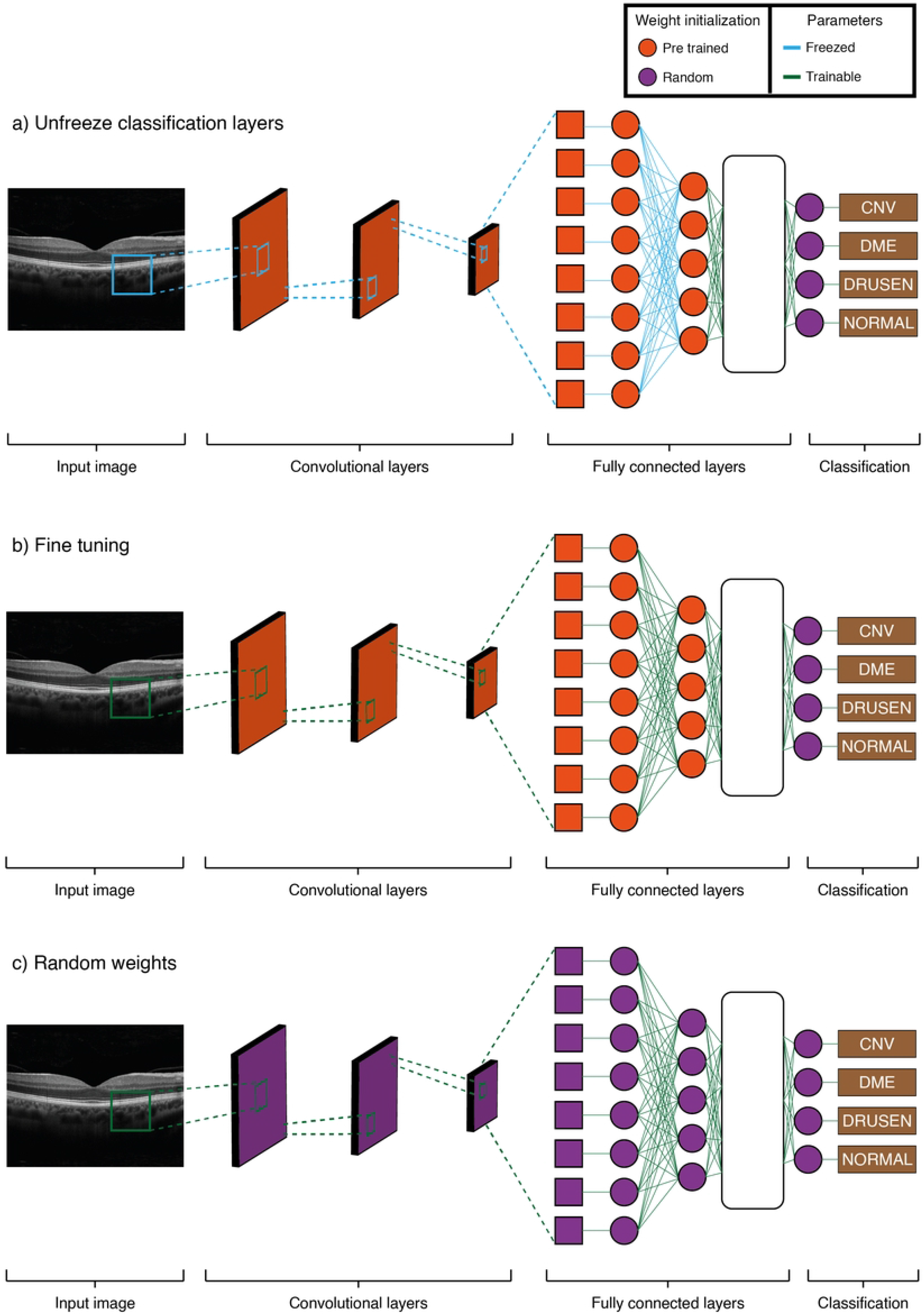
Differentiated Training Methodologies for CNN Models: a) Unfreezing Classification Layer utilizes pre-trained network parameters, freezing all except the final layers for targeted training. b) Fine-Tuning permits updates to all layers of a pre-trained network, integrating newly initialized layers. c) Training from scratch establishes a fully trainable network with no transfer learning, starting with a random initialization of all layers.

**Table 5:**
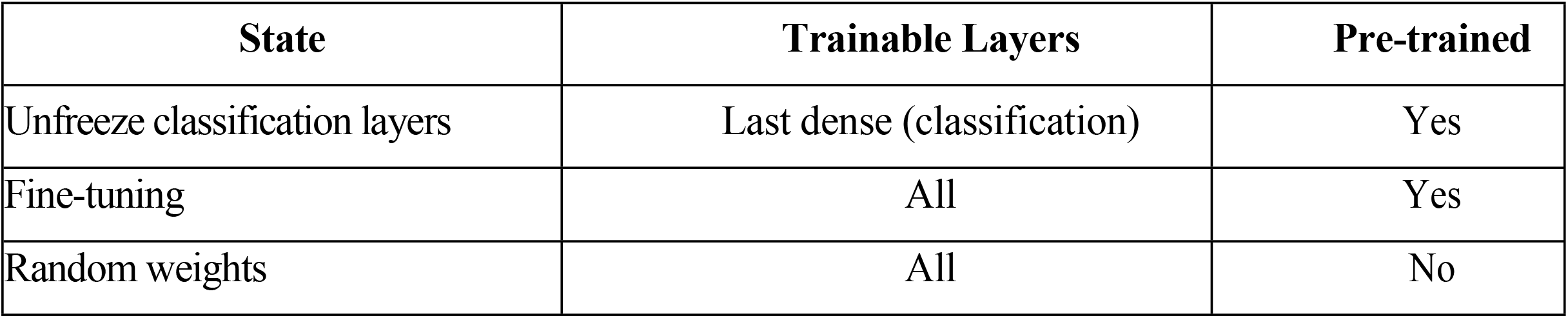
Overview of CNN model training approaches highlighting trainable layers and use of pre-training.

**Algorithm 1:**
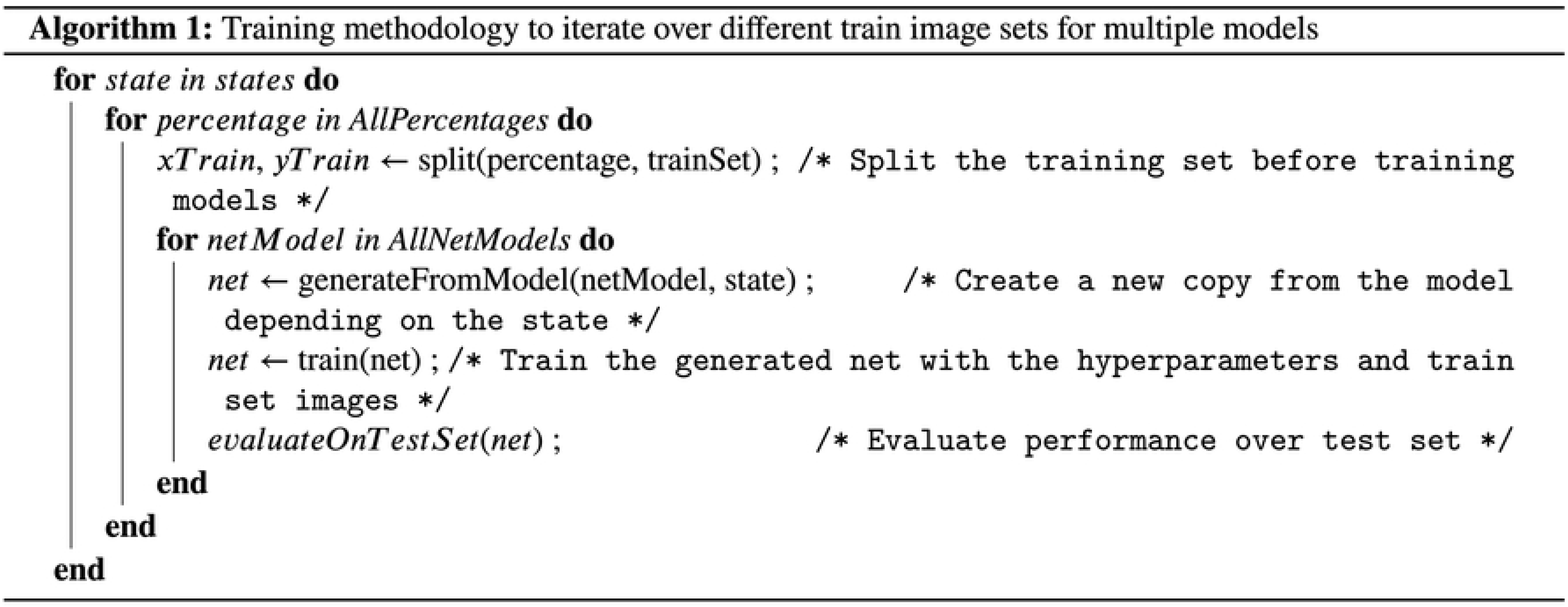
Iterative training methodology across various image sets for multiple model evaluation.

#### 2.2.4. **Performance Metrics**

To assess the performance of models with differing volumes of input data, standard classification metrics were utilized. These metrics provide insights into the accuracy and effectiveness of each model’s predictive capabilities.

##### Confusion Matrix

The confusion matrix is an integral tool in classification tasks, comparing actual to predicted labels. It’s particularly important for binary classification, with four primary components: true positives (TP), true negatives (TN), false positives (FP), and false negatives (FN). These components help to identify both correct predictions and various types of errors (see Figure 3). For problems involving more than two classes, the matrix expands to cover all category permutations, aiding in the detailed analysis of model predictions.

**Figure 3:**
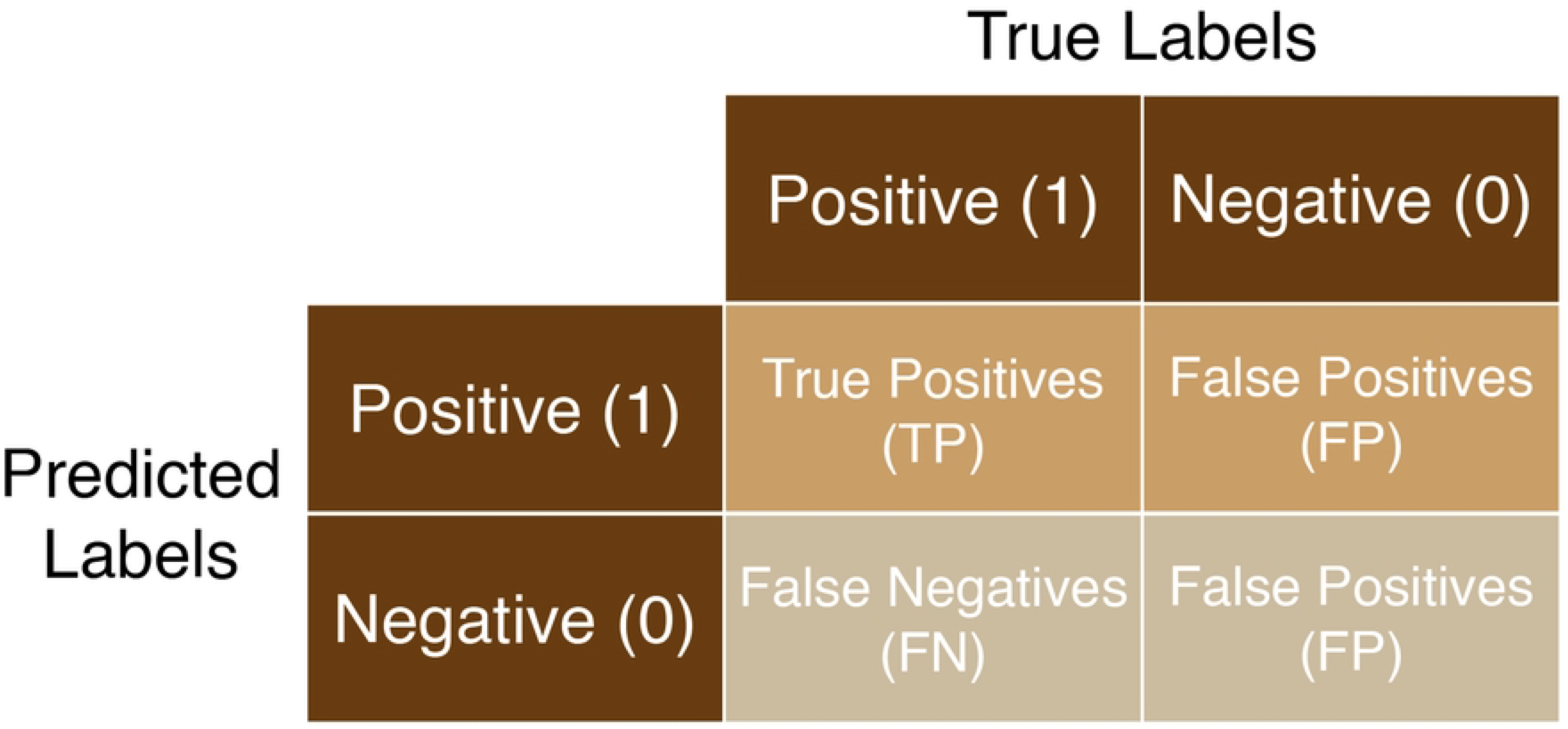
Confusion matrix for a binary classification problem. The matrix diagonal corresponds to the correct predictions, whereas any other value corresponds to *confusion* between the predicted and true labels.

##### Accuracy

Accuracy is a straightforward metric that calculates the proportion of correctly identified instances in relation to all the predictions made by the classification model for OCT image datasets (see equation 4.1).

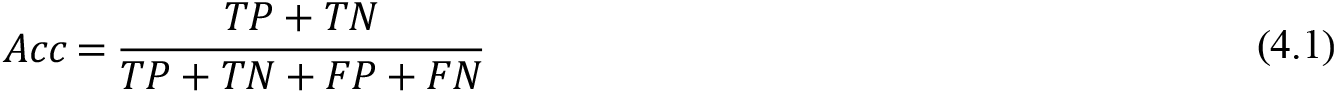

##### IoU (Intersection over Union)*

IoU is a metric that evaluates the proportion of overlap between the predicted positive cases and the actual positive cases, excluding the true negatives. It’s a measure of the accuracy of the model’s predictions. (see equation 4.2).

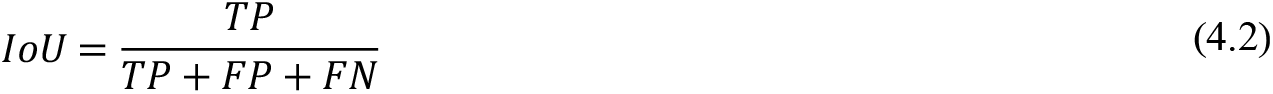

##### Recall (sensitivity)*

This metric assesses how effectively the model can identify correct instances of a specific class within the dataset (see equation 4.3).

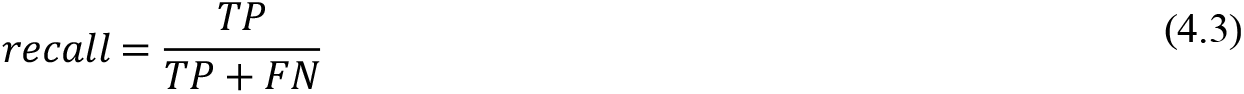

##### Precision*

Precision measures the accuracy of the model’s positive predictions, specifically the proportion of actual positives among all instances classified as positive (see equation 4.4).

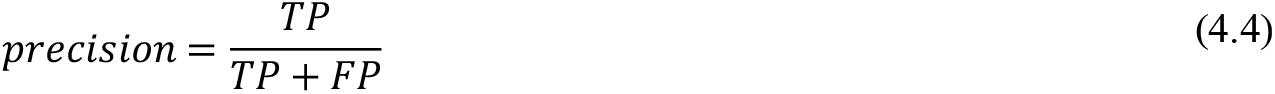

##### F1-Score*

This metric balances precision, which is the correctness of positive predictions, and sensitivity, the model’s ability to correctly identify all relevant instances (see equation 4.5).

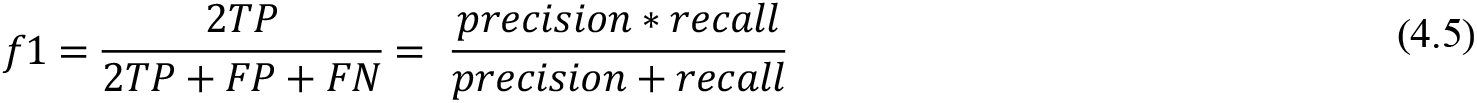

##### Cohen Kappa

The Cohen Kappa coefficient is a metric for assessing the accuracy of a model’s predictions, offering a measure of reliability that is particularly useful in contexts with imbalanced datasets (see equation 4.6).

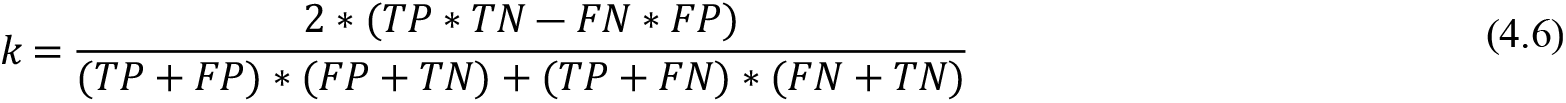

##### AUC (Area Under the Receiver Operating Characteristic Curve)

The AUC represents the area under the ROC curve, which illustrates a model’s capacity to distinguish between classes at various threshold settings. It reflects the balance between correctly identifying true positives and avoiding false positives.

In multiclass classification problems, such as identifying the four categories DME, Drusen, CNV, and Normal metrics marked with an asterisk (*) use a weighted average strategy. This involves comparing each label individually, then adjusting the metric based on the prevalence of each label in the dataset, and finally averaging the results across all labels.

## 3. **Results**

Table 1 provides a comprehensive summary of studies employing machine learning and deep learning techniques for the analysis of Optical Coherence Tomography (OCT) images, covering the period from 2014 to 2024. It details the diverse architectures utilized, including SVM, VGG16, AlexNet, GoogLeNet, and InceptionV3, among others, many of which are pre-trained on extensive datasets. The table categorizes each study by the datasets used, the ocular features classified (such as Diabetic Macular Edema (DME), Choroidal Neovascularization (CNV), and Drusen), and the performance metrics reported (including accuracy, sensitivity, specificity, precision, F1-score, and AUC). These studies collectively highlight significant progress in the field, showcasing the enhanced accuracy and efficiency in classifying ocular pathologies through OCT image analysis. Notwithstanding the impressive accuracy, often exceeding 99%, achieved by these studies in classifying retinal pathologies using OCT images, the impact of model learning capacity relative to data volume remains underexplored.

The training of all models was conducted using Python with the Keras library on TensorFlow version 2.4.1. These operations were carried out on a server equipped with an Intel(R) Xeon(R) Bronze 3204 six-core 64-bit processor, 256GB of DDR4 RAM, and an NVIDIA RTX 8000 GPU with 48GB of DDR5 VRAM.

### 3.1. **Metrics and Performance**

The metrics results, as presented in the figures 4-7, illustrate the performance of various training methodologies. Figure 4 indicates that a maximum of thirty epochs sufficed to optimize accuracy without overfitting. Figure 5 contrasts the accuracy and F1 score against the volume of training images, noting performance gains in models trained with random weights as the image count increased.

**Figure 4:**
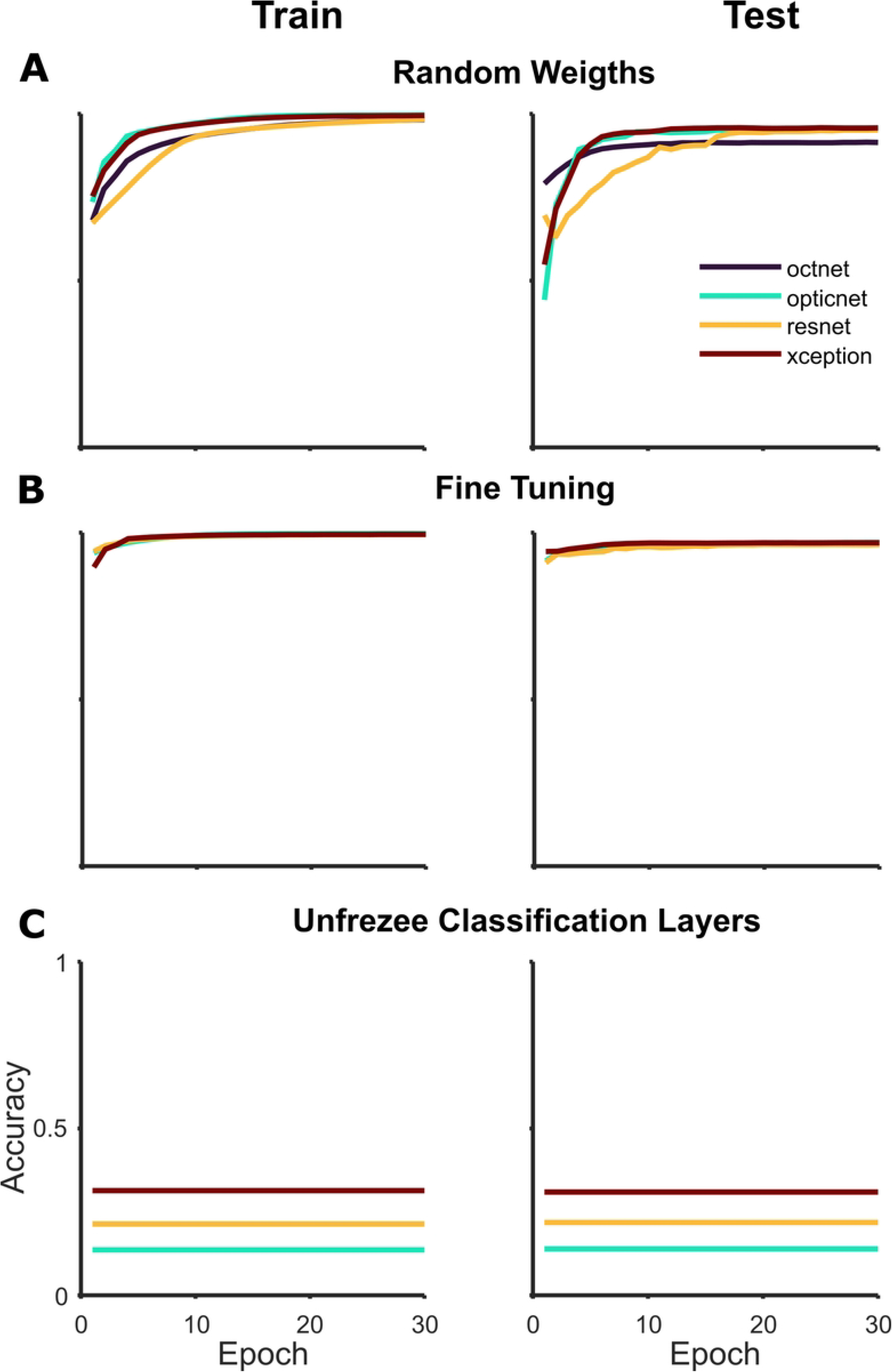
Training Accuracy over Epochs on three training scenaries: This graph displays training accuracy over 30 epochs for different models under three training scenarios: (A) using random weights, (B) applying fine-tuning, and (C) unfreezing classification layers. Each curve traces a model’s learning progression, with epochs on the horizontal axis and accuracy on the vertical.

**Figure 5:**
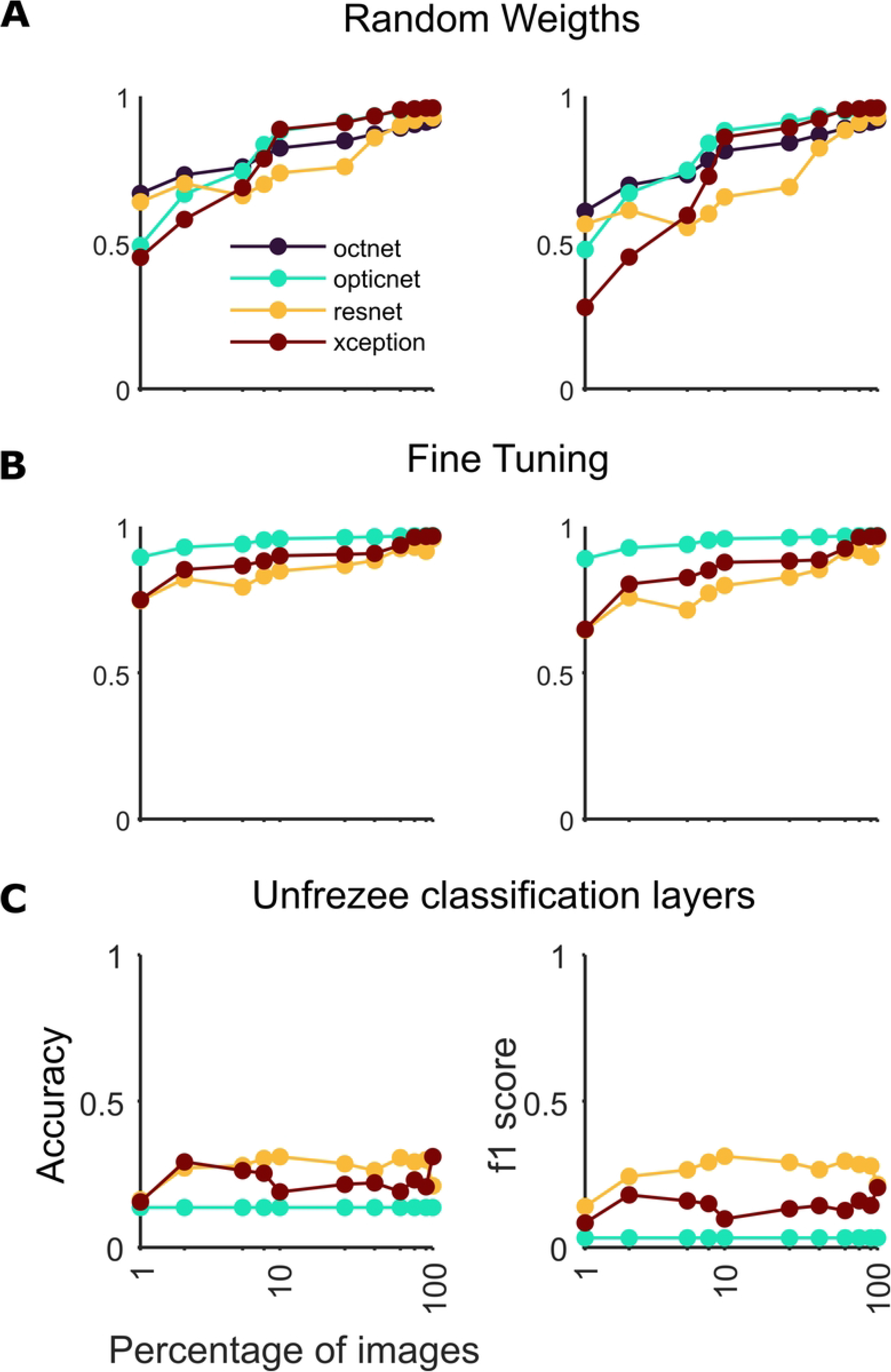
Curves depicting the test set performance across different training set sizes in logarithmic scale: (A) showcases accuracy and F1-score for models trained with random weights; (B) presents these metrics for models using the fine-tuning approach, excluding OctNET; and (C) illustrates outcomes for models with only the classification layers trained.

OpticNet consistently outperformed other models with both random weights and fine-tuning, irrespective of training set size. ResNet-50, while underperforming with random weights, was surpassed by Xception in the fine-tuning scenario. OctNet, lacking pre-training, excelled only with random weights under OpticNet. All models showed poor performance when only the classification layers were unfrozen, equivalent to a random guess. Training times, visualized in Figure 6, correlated with architecture complexity: larger models with more parameters demanded longer training periods, showing an exponential relationship with size.

**Figure 6:**
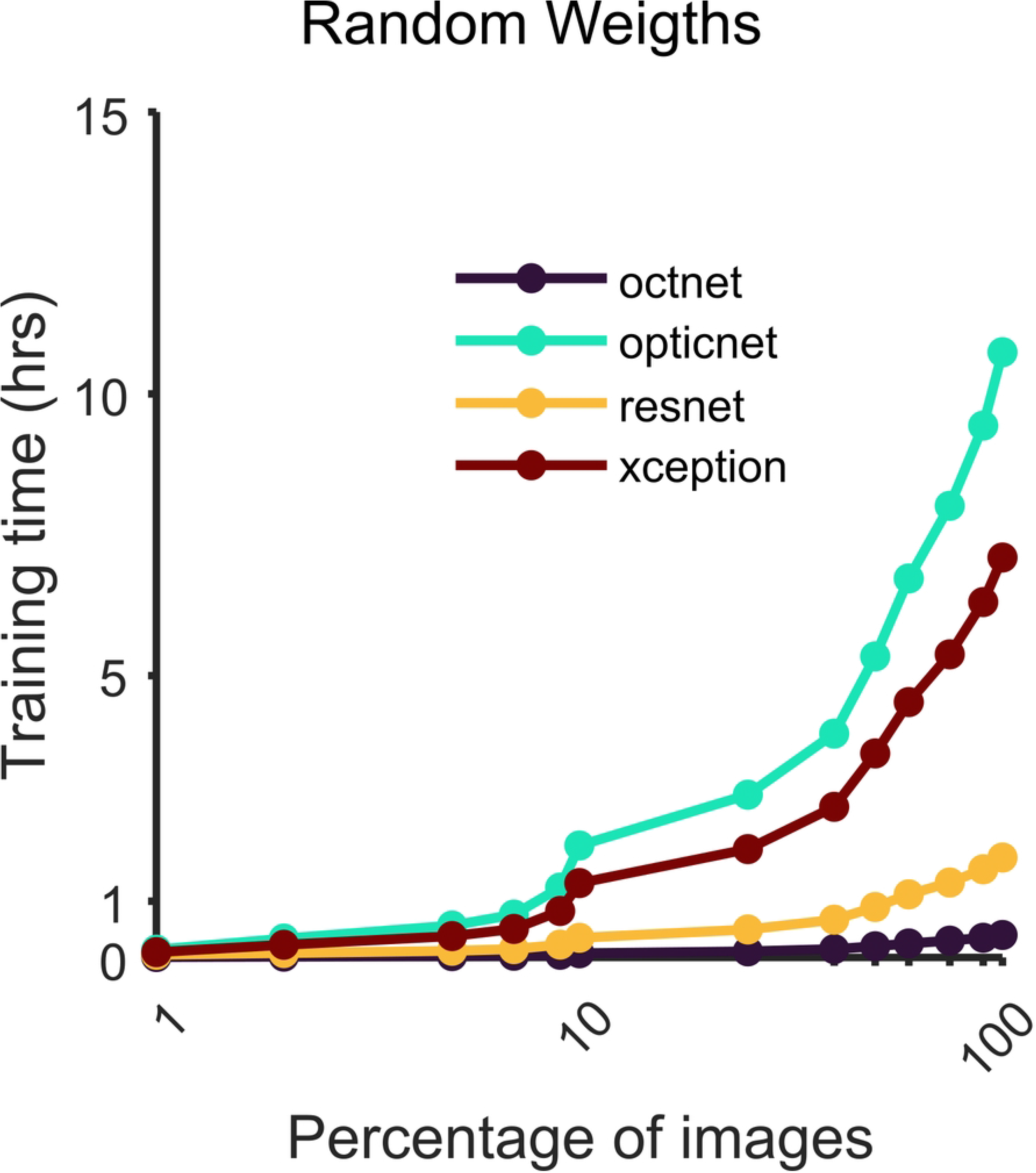
Training times in logarithmic scale for each model along the train set size percentage in logarithmic scale in x-axis for all the proposed models.

### 3.2. **Performance Comparison**

Figure 7 contrasts the confusion matrices and ROC curves for the best and worst-performing models in detecting DME against other categories. It compares the performance of ResNet-50 trained on 1% of the data to OpticNet trained on the full dataset. OpticNet achieved near-perfect accuracy, while ResNet-50’s accuracy was significantly lower, with widespread misclassifications across categories. Notably, ResNet-50 often confused DRUSEN for CNV and mislabeled NORMAL images as DRUSEN. Table 6 presents a comparative analysis of the performance of ResNet-50, Xception, and OpticNet models across varying training conditions and dataset sizes. Performance metrics such as accuracy, Intersection over Union (IoU), recall, precision, F1-score, Cohen Kappa, and Area Under the Curve (AUC) offer insights into each model’s capabilities across different training methodologies, namely unfreezing classification layers, fine-tuning, and employing random weights. The analysis reveals significant performance enhancements correlated with increased data volume and the effectiveness of fine-tuning in optimizing model accuracy. OctNET’s results are presented solely for the random weights approach, showcasing a significant improvement in performance as training data volume increases.

**Figure 7:**
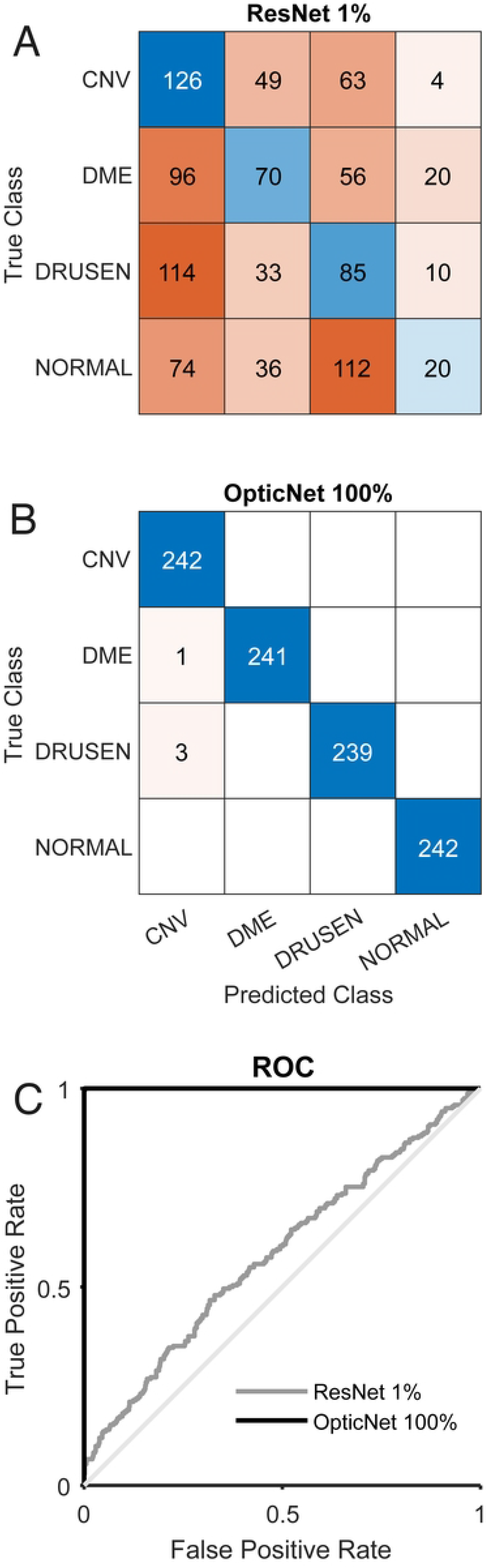
Overall results comparison. A. Confusion matrix for ResNet-50 with unfroze last classification layers trained with just 1% of the complete training set. B. Confusion matrix of the OpticNet model trained with the complete training set (blank squares denote a zero value). C. ROC curve of the above models (ResNet with 1% training set and OpticNet with 100%).

**Table 6:** Comparative performance metrics of selected CNN models across three different training methodologies.

| Model | Train images | Train methodology | Accuracy | IoU | Recall | Precision | F1-score | Cohen-Kappa | AUC |
| --- | --- | --- | --- | --- | --- | --- | --- | --- | --- |
| ResNet- 50 | 835 | Unfreeze classification | 0.311 | 0.171 | 0.311 | 0.329 | 0.288 | 0.081 | 0.545 |
|  |  | Fine-tuning | 0.499 | 0.320 | 0.499 | 0.321 | 0.369 | 0.332 | 0.945 |
|  |  | Random weights | 0.250 | 0.063 | 0.250 | 0.063 | 0.100 | 0.000 | 0.506 |
|  | 8,348 | Unfreeze classification | 0.263 | 0.125 | 0.263 | 0.262 | 0.212 | 0.018 | 0.542 |
|  |  | Fine-tuning | 0.983 | 0.968 | 0.983 | 0.984 | 0.983 | 0.978 | 0.992 |
|  |  | Random weights | 0.974 | 0.950 | 0.974 | 0.976 | 0.974 | 0.966 | 0.994 |
|  | 83,484 | Unfreeze classification | 0.222 | 0.106 | 0.222 | 0.189 | 0.185 | -0.037 | 0.435 |
|  |  | Fine-tuning | **0.999** | **0.998** | **0.999** | **0.999** | **0.999** | **0.999** | **1.000** |
|  |  | Random weights | **0.999** | **0.998** | **0.999** | **0.999** | **0.999** | **0.999** | **1.000** |
| Xception | 835 | Unfreeze classification | 0.248 | 0.073 | 0.248 | 0.157 | 0.120 | -0.003 | 0.522 |
|  |  | Fine-tuning | 0.500 | 0.325 | 0.500 | 0.325 | 0.371 | 0.333 | 0.925 |
|  |  | Random weights | 0.264 | 0.078 | 0.264 | 0.165 | 0.128 | 0.019 | 0.516 |
|  | 8,348 | Unfreeze classification | 0.253 | 0.076 | 0.253 | 0.211 | 0.127 | 0.004 | 0.508 |
|  |  | Fine-tuning | **0.999** | **0.998** | **0.999** | **0.999** | **0.999** | **0.999** | **1.000** |
|  |  | Random weights | 0.996 | 0.992 | 0.996 | 0.996 | 0.996 | 0.994 | 1.000 |
|  | 83,484 | Unfreeze classification | 0.253 | 0.076 | 0.253 | 0.211 | 0.127 | 0.004 | 0.507 |
|  |  | Fine-tuning | **0.999** | **0.998** | **0.999** | **0.999** | **0.999** | **0.999** | **1.000** |
|  |  | Random weights | 0.998 | 0.996 | 0.998 | 0.998 | 0.998 | 0.997 | 1.000 |
| OpticNet | 835 | Unfreeze classification | 0.250 | 0.063 | 0.250 | 0.063 | 0.100 | 0.000 | 0.496 |
|  |  | Fine-tuning | 0.978 | 0.958 | 0.978 | 0.980 | 0.978 | 0.971 | 1.000 |
|  |  | Random weights | 0.564 | 0.381 | 0.564 | 0.646 | 0.533 | 0.419 | 0.846 |
|  | 8,348 | Unfreeze classification | 0.250 | 0.063 | 0.250 | 0.063 | 0.100 | 0.000 | 0.496 |
|  |  | Fine-tuning | 0.996 | 0.992 | 0.996 | 0.996 | 0.996 | 0.994 | 1.000 |
|  |  | Random weights | **0.999** | **0.998** | **0.999** | **0.999** | **0.999** | **0.999** | **1.000** |
|  | 83,484 | Unfreeze classification | 0.250 | 0.063 | 0.250 | 0.063 | 0.100 | 0.000 | 0.496 |
|  |  | Fine-tuning | 0.996 | 0.992 | 0.996 | 0.996 | 0.996 | 0.994 | 1.000 |
|  |  | Random weights | **0.999** | **0.998** | **0.999** | **0.999** | **0.999** | **0.999** | **1.000** |
| OctNET | 835 | Random weights | 0.588 | 0.380 | 0.588 | 0.646 | 0.511 | 0.450 | 0.872 |
|  | 8,348 | Random weights | 0.897 | 0.812 | 0.897 | 0.908 | 0.896 | 0.862 | 0.991 |
|  | 83,484 | Random weights | 0.987 | 0.974 | 0.987 | 0.987 | 0.987 | 0.982 | 1.000 |

## 4. **Discussion**

Recent studies have consistently demonstrated high accuracy, often exceeding 99%, with differences emerging at the third decimal place (see Table 1). Despite the increasing number of publications suggesting modifications to existing models or the development of new ones, these studies tend to prioritize complexity over efficiency, frequently overlooking the computational cost and training time. There is also a lack of consideration for training these models with subsets of the available data, rather than the full dataset.

In this research, the models implemented and trained have surpassed all those reviewed in the systematic analysis, achieving up to 99.9% accuracy and recall. The exception is OctNET, which attained 98% precision, deviating slightly from the original paper. Such discrepancies might stem from variations in the training hyperparameters or how the dataset was divided. Nonetheless, the difference in reported performance is marginal, under 1%. Notably, OpticNet’s performance with random weights and a mere 10% of the training set surpassed any other model trained on the Kermany dataset, which typically utilized the full dataset.

Fine-tuning is the leading strategy, significantly outperforming the unfreezing classification layers approach [23], [34]. This underperformance is attributed to a constant learning rate, contrary to the recommended practice of adjusting it during retraining to encourage convergence. The inadequacy of the unfreezing method is highlighted by results comparable to a random classifier, as seen in Figure 6. On the other hand, the random weights approach shows promise with larger datasets, achieving comparable accuracy to fine-tuning beyond a certain data volume.

The analysis indicates that larger training sets typically improve model performance. However, a key finding is that there’s a threshold approximately 10% of the full training set, or 8,348 image where models fine-tuned on this subset already achieve above 98% accuracy. With random weights, models reach at least 97% accuracy, except for OctNET, which requires around 41,000 images to hit the 97% mark. For the Xception and OpticNet models, fine-tuning on as few as 6,261 and 4,174 images, respectively, suffices to achieve 99% accuracy. Generally, with fine-tuning or random weights, models attain over 99% accuracy starting from training sets with at least 6,261 images.

The growth of training time with an increase in training set size is exponentially greater across all model architectures. With fewer parameters to train, there’s a significant reduction in computational time. OctNET stands out for its exceptionally rapid training time, which is an order of magnitude faster than other models like OpticNet or Xception. This efficiency becomes critical for models requiring frequent updates with new data, as the time difference between mere minutes for OctNET and several hours for others is a crucial consideration for continuous retraining and deployment.

OpticNet delivered the highest performance, likely due to its design, which is specialized for OCT image classification. Despite this, it shared similar challenges with other models when only the classification layers were unfrozen. In fine-tuning scenarios, OpticNet’s advantage is more pronounced, possibly because it was pre-trained on a relevant dataset. Interestingly, OctNET’s performance was on par with pre-trained models, challenging the expectation that pre-training should lead to quicker improvements with increasing dataset size. This could indicate that the pre-trained features were not as advantageous for OCT classification or that OCT images do not require complex models for high accuracy.

## 5. **Conclusion**

This study performed a systematic review to identify research papers where machine learning algorithms were applied to diagnose retinal pathologies from OCT images. A selection of high-ranking models from the literature was tested using widely used datasets, and three distinct training strategies were evaluated. This comparison aimed to determine the minimum dataset size required for robust model training. Furthermore, the study analyzed the trade-off between model performance metrics and training time to determine the most practical models for real-world application, with OpticNet emerging as the most effective in several training contexts.

The systematic review revealed that prior research did not comprehensively assess the impact of training set size, typically adhering to the maxim “more data yields better results”. This study challenges that notion, demonstrating certain CNN architectures can reach optimal performance with only about 10% of the full dataset. It further illustrates that a model with fewer parameters less than 6% of the total can achieve comparable outcomes, significantly reducing both computational demands and hardware needs. Moreover, models pre-trained on specialized datasets like OpticNet with the Kermany dataset are immediately effective for specific tasks, such as detecting retinal pathologies. Conversely, models like ResNet and Xception, pre-trained on the more generalized ImageNet, might require additional tuning for specialized medical imaging tasks. This situation demonstrates why models with random initial weights can sometimes match or exceed the performance of those with specific pre-training, highlighting the importance of dataset relevance over general pre-training.

## Code Availability

All the code has been made available in the following GitHub repository: https://github.com/HpcDataLab/DL_ENESJ_IMO/

## Author Contributions

The study’s design and conceptualization were the collective work of Walter Hauri-Rosales (W.H.R.), Oswaldo Pérez (O.P.), Marlon Garcia-Roa (M.G.R.), Ellery López-Star (E.L.S.), and Ulises Olivares-Pinto (U.O.P.). The training of the models was executed by W.H.R., O.P., and U.O.P., who also conducted the data analysis, which included statistical assessments. The manuscript was collaboratively written, reviewed, and edited by all the authors, who have given their approval for the final version to be submitted.

## Data Availability

https://github.com/HpcDataLab/DL_ENESJ_IMO/

## Acknowledgments

The authors thank the Instituto Mexicano de Oftanmología (IMO) IAP for providing support. Recognition is also due to the financial support received from CONACyT (CF:191982) (UOP) and the UNAM-DGAPA-PAPIIT “TA101022” (UOP) and “TA101124” (UOP).

## Statement

During the preparation of this work, the authors improved the readability and clarity of the text using commercial grammar tools. After utilizing these tools, the author(s) reviewed and edited the content as needed and take full responsibility for the content of the publication.

